# An Early and Preliminary Assessment of the Clinical Severity of the Emerging SARS-CoV-2 Omicron Variants in Maharashtra, India

**DOI:** 10.1101/2022.09.07.22279665

**Authors:** Rajesh Karyakarte, Rashmita Das, Nyabom Taji, Sushma Yanamandra, Smriti Shende, Suvarna Joshi, Bhagyashree Karekar, Reshma Bawale, Rahul Tiwari, Madhuri Jadhav, Shivani Sakalkar, Geetanjali Chaudhari, Srushti Rane, Jeanne Agarasen, Praveena Pillai, Sonali Dudhate, Priyanka Chandankhede, Rutika Labhshetwar, Yogita Gadiyal, Mansi Rajmane, Savita Mukade, Preeti Kulkarni

## Abstract

**Background:** The SARS-CoV-2 Omicron variants BA.2.74, BA.2.75 and BA.2.76 have appeared recently in India and have already spread to over 40 countries. They have acquired additional mutations in their spike protein compared to BA.2, branching away on the SARS-CoV-2 phylogenetic tree. These added mutations, over and above those of the parental BA.2 variant, have raised concerns about the impact on viral pathogenicity, transmissibility, and immune evasion properties of the new variants.

**Material and Methods:** A total of 990 RT-PCR positive SARS-CoV-2 samples, with a cycle threshold value (Ct) less than 25, were processed for SARS-CoV-2 whole genome sequencing between 3rd June 2022 to 7th August 2022. All corresponding demographic and clinical data were recorded and analyzed using Microsoft® Excel.

**Results:** Out of 990 samples sequenced, BA.2.75 (23.03%) was the predominant Omicron sublineage, followed by BA.2.38 (21.01%), BA.5 (9.70%), BA.2 (9.09%), BA.2.74 (8.89%) and BA.2.76 (5.56%). A total of 228 cases of BA.2.74, BA.2.75 and BA.2.76 were contacted by telephone, of which 215 (94.30%) were symptomatic with mild symptoms, and 13 (5.70%) had no symptoms. Fever (82.02%) was the most common symptom, followed by cough (49.12%), cold (35.97%), fatigue (27.19%), headache (21.05%) and myalgia (20.61%). Of the 228 cases, 195 (85.53%) cases recovered at home, and 33 (14.47%) required institutional quarantine. Recovery with conservative treatment was observed in 92.98% of cases, while 4.83% required additional oxygen therapy. Only 03 (1.32%) cases had poor outcomes resulting in death, and the remaining 225 (98.68%) had a good outcome. Among the 228 cases, 219 (96.05%) cases were vaccinated with COVID-19 vaccine; of these 72.60% had received both doses, 26.03% had also received the precautionary booster dose, while 1.37% were incompletely vaccinated with a single dose of vaccine.

**Conclusion:** The current study indicates that the three BA.2 sublineages are causing mild disease in India. However, BA.2.75 has key mutations that are notable for accelerated growth and transmission and require close and effective monitoring.

## 1. INTRODUCTION

Since the first case of Severe Acute Respiratory Syndrome Coronavirus (SARS-CoV-2) in March 2020, India has witnessed three pandemic waves. Delta (B.1.617.2) and its sublineages (AY. *) caused the second wave, and Omicron (B.1.1.529) and its sublineages (BA.1 and BA.2) are driving the third wave. (1)

The B.1.1.529 was first reported from South Africa on 24^th^ November 2021 (2) and was designated as a Variant of Concern (VoC) on 26^th^ November 2021 by World Health Organisation. By late January 2022, the Omicron variant had been identified in 171 countries across all six WHO regions. Due to its substantial growth advantage over Delta, the Omicron variant rapidly replaced the Delta variant globally. Initially, the Omicron variant (B.1.1.529) comprised three sister lineages, B.1.1.529.1 (BA.1), B.1.1.529.2 (BA.2) and BA.1.1.529.3 (BA.3). Characteristic constellation of mutations, particularly 26-32 mutations in its spike protein, have been responsible for its increased transmissibility and its ability to evade immunity established by natural infection or vaccination. (3)

India witnessed its third wave from late December 2021 to late February 2022 (4), with BA.1 and BA.2 dominating the early and the latter half of the wave, respectively (3). Since then, the Omicron variant has evolved continuously to give rise to BA.4/ BA.5 in South Africa (5) and BA.2.12.1 in the USA (6), causing global outbreaks. Unlike the Delta variant, Omicron caused less severe disease and decreased hospitalization rates and deaths. However, unvaccinated people with advanced age and underlying disease conditions suffered from severe disease. (7)

After the waning of the third wave, India saw a surge in COVID-19 cases from the fourth week of May 2022 (4). On sequencing, these variants were characterized as BA.2 by Pangolin. However, the predominance of BA.2 after the waning of the third COVID-19 wave was unexplainable. This issue was discussed in detail on the GitHub repository, covlineages/pango-designation, a repository for suggesting new lineages that should be added to the current scheme for naming the SARS-CoV-2 virus. Subsequently, the Indian isolates of BA.2 were further classified into sub-lineages BA.2.74 (Issue #775) (8), BA.2.75 (Issue #773) (9) and BA.2.76 (Issue #787) (10). The BA.2.75 subvariant has been classified as an Omicron subvariant under monitoring by the World Health Organization (WHO) due to its increasing numbers in India and its identification in dozen countries, including Nepal, Singapore, Martinique, China, New Zealand, Cambodia and Indonesia. (11) Only three months since its designation, BA.2.75 subvariant has acquired several mutations to give rise to new sublineages (BA.2.75.1, BA.2.75.2, BA.2.75.3, BA.2.75.4, BA.2.75.6, BL.1, BM.1 and BN.1) that can compete with the circulating lineages. (12)

With the emergence of new variants, data on the clinical severity of the disease caused by them are essential to guide public health planning and response. Therefore, the current study aimed to describe the severity and clinical presentation of these emerging SARS-CoV-2 variants sequenced at Byramjee Jeejeebhoy Government Medical College, Pune, Maharashtra.

## 2. MATERIAL AND METHODS

This study was carried out at the Department of Microbiology of the Byramjee Jeejeebhoy Government Medical College (BJGMC), Pune, Maharashtra. It was part of the Indian SARS-CoV-2 Genomics Consortium (INSACOG) sequencing activity in Maharashtra. INSACOG is a Pan-India network of 54 laboratories to monitor the genomic variations in the SARS-CoV-2 virus and to study the linkages between the variants and epidemiological trends.

The study falls within the research activities approved by the BJGMC Institutional Ethics Committee, Pune, Maharashtra, India.

### 2.1. Sample Acquisition

The Department of Microbiology of BJGMC, Pune, is the coordinating laboratory for the surveillance of the SARS-CoV-2 virus in the community in Maharashtra. It receives samples from its attached hospital and various RTPCR testing laboratories across Maharashtra for SARS-CoV-2 whole genome sequencing. Nasopharyngeal samples collected in VTM, between 3rd June 2022 to 7th August 2022 with a cycle threshold (Ct) value of less than 25, were transported to the laboratory, maintaining a cold chain at 2 to 8°C. The samples were stored at −80°C until further processing.

The viral RNA was extracted using the MagMax™ Viral/Pathogen Nucleic Acid Isolation Kit (Thermofisher Scientific Inc., Waltham, US), following the manufacturer’s instructions. The RNA samples were processed for SARS-CoV-2 whole genome sequencing, and its analysis was done at the Centre of Excellence for Genomics, Department of Microbiology, BJGMC, Pune.

### 2.2. Library preparation, next-generation sequencing, and lineage analysis

Libraries for Covid-19 were prepared using Rapid Barcoding and Midnight RT-PCR Expansion kits (Midnight protocol) (Oxford Nanopore Technologies (ONT), Littlemore, United Kingdom). Sequencing was conducted on the R9.4 flow cell by ONT using a GridION sequencer (ONT, United Kingdom). Primary data acquisition was performed using MinKNOW, version 22.05.7, the operating software that operates nanopore sequencing devices. Basecalling was performed using Guppy, version 6.1.5, in fast basecalling mode. The data was further processed using the wf-ARTIC workflow, a repository that contains the nextflow workflow for running the ARTIC SARS-CoV-2 workflow on multiplexed GridION runs, installed in MinKNOW software. Lineage identification was carried out using Pangolin, version 4.1.1, pangolin-data version 1.12 and University of California Santa Cruz (UCSC) Genome Browser UShER: Ultrafast Sample placement on Existing tree. The clade analysis was done using the Nextclade software, version 2.3.0.

### 2.3. Data Collection

A set of individual-level data was obtained, corresponding to the samples received from various RT-PCR laboratories sending samples for sequencing. Each sample’s unique identification number (ICMR ID) was also recorded. Additional information on the presence of any symptoms, hospitalization, treatment, comorbidities and vaccination status was collected via a telephonic interview with each patient.

### 2.4. Statistical analysis

All demographic and clinical data were recorded and analyzed using Microsoft® Excel.

## 3. RESULTS

Between 3^rd^ June 2022 and 7^th^ August 2022, 990 RT-PCR positive SARS-CoV-2 samples were processed and sequenced at BJGMC, Pune. The study population included cases from all age groups with a median age of 36 years. The male-to-female ratio was 1.39:1. **Table 1** shows the geographical distribution of the sequenced samples.

**Table 1:** Geographical Distribution of 990 RT-PCR positive SARS-CoV-2 samples

| Geographical Area | Number (%) |
| --- | --- |
| Pune | 792<br>(80%) |
| Mumbai | 143<br>(14.44%) |
| Solapur | 27<br>(2.73%) |
| Latur | 16<br>(1.62%) |
| Jalgaon | 7<br>(0.71%) |
| Sindhudurg | 5<br>(0.50%) |
| **Grand Total** | **990**<br>**(100%)** |

Of these 990 samples sequenced, BA.2.75 (23.03%) was the predominant Omicron sublineage followed by BA.2.38 (21.01%), BA.5 (9.70%), BA.2 (9.09%), BA.2.74 (8.89%) and BA.2.76 (5.56%) **(Table 2)**.

**Table 2:**
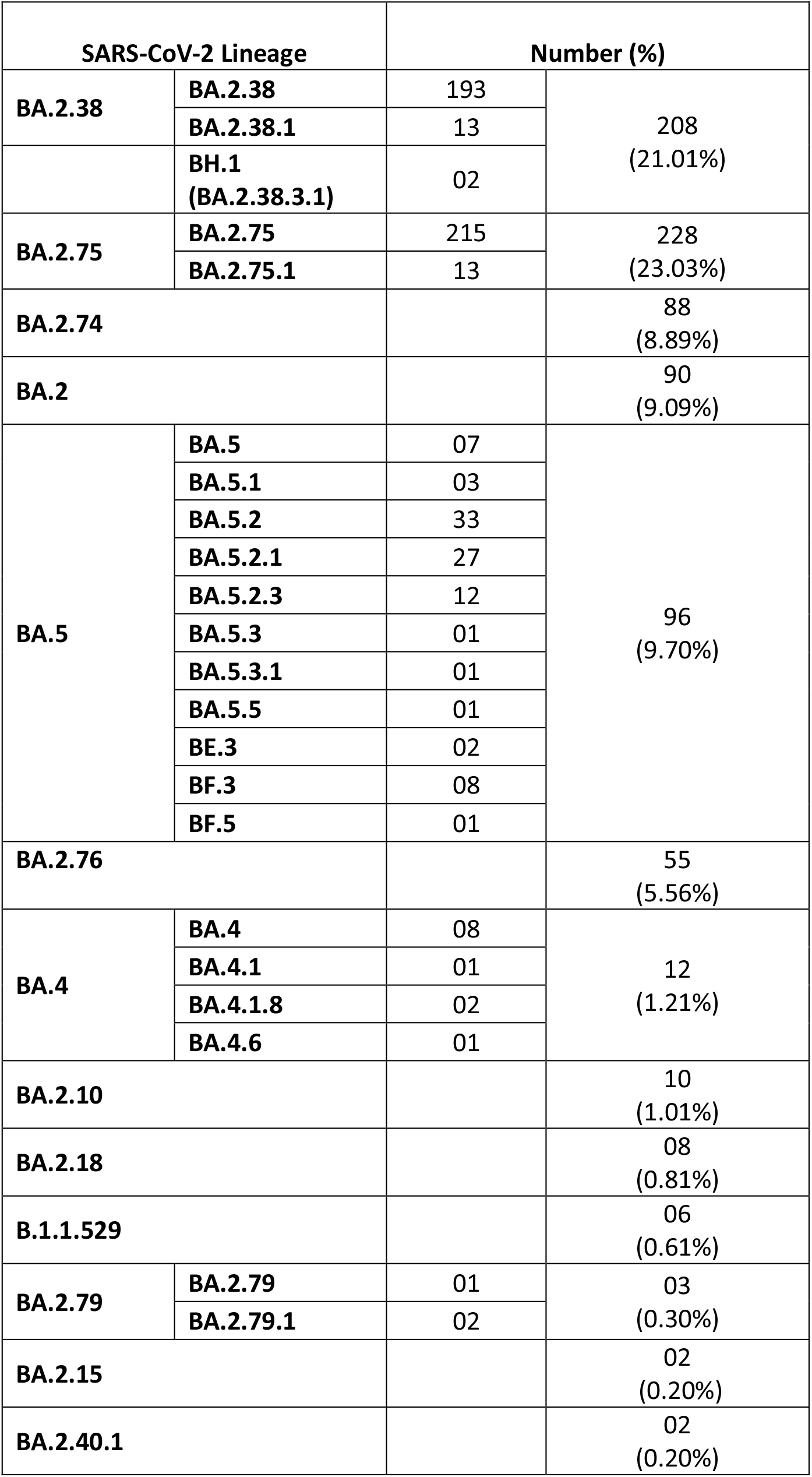
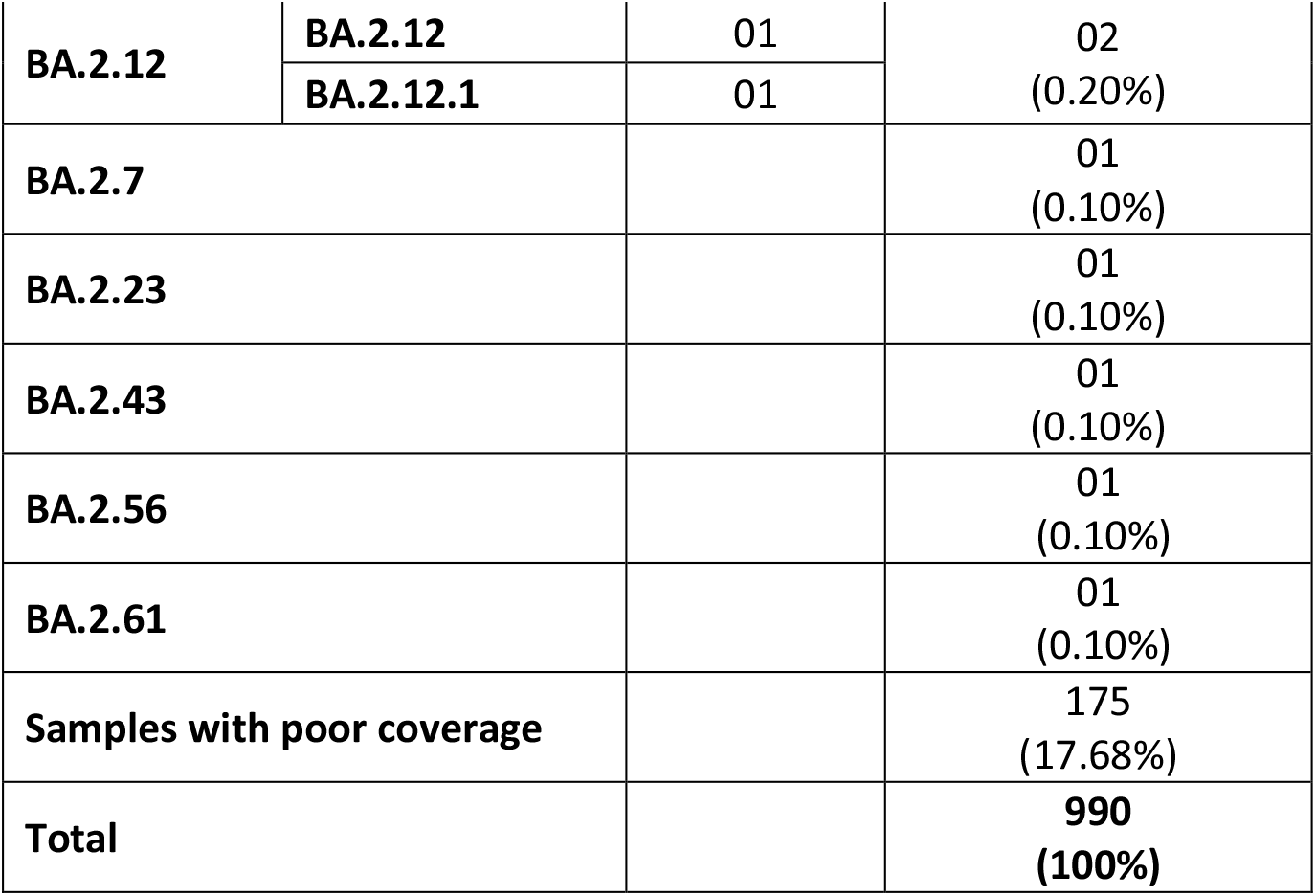
Variant Distribution among the 990 SARS-CoV-2 samples sequenced

The distribution of variants to date of sample collection is shown in **Figure 1**.

**Figure 1:**
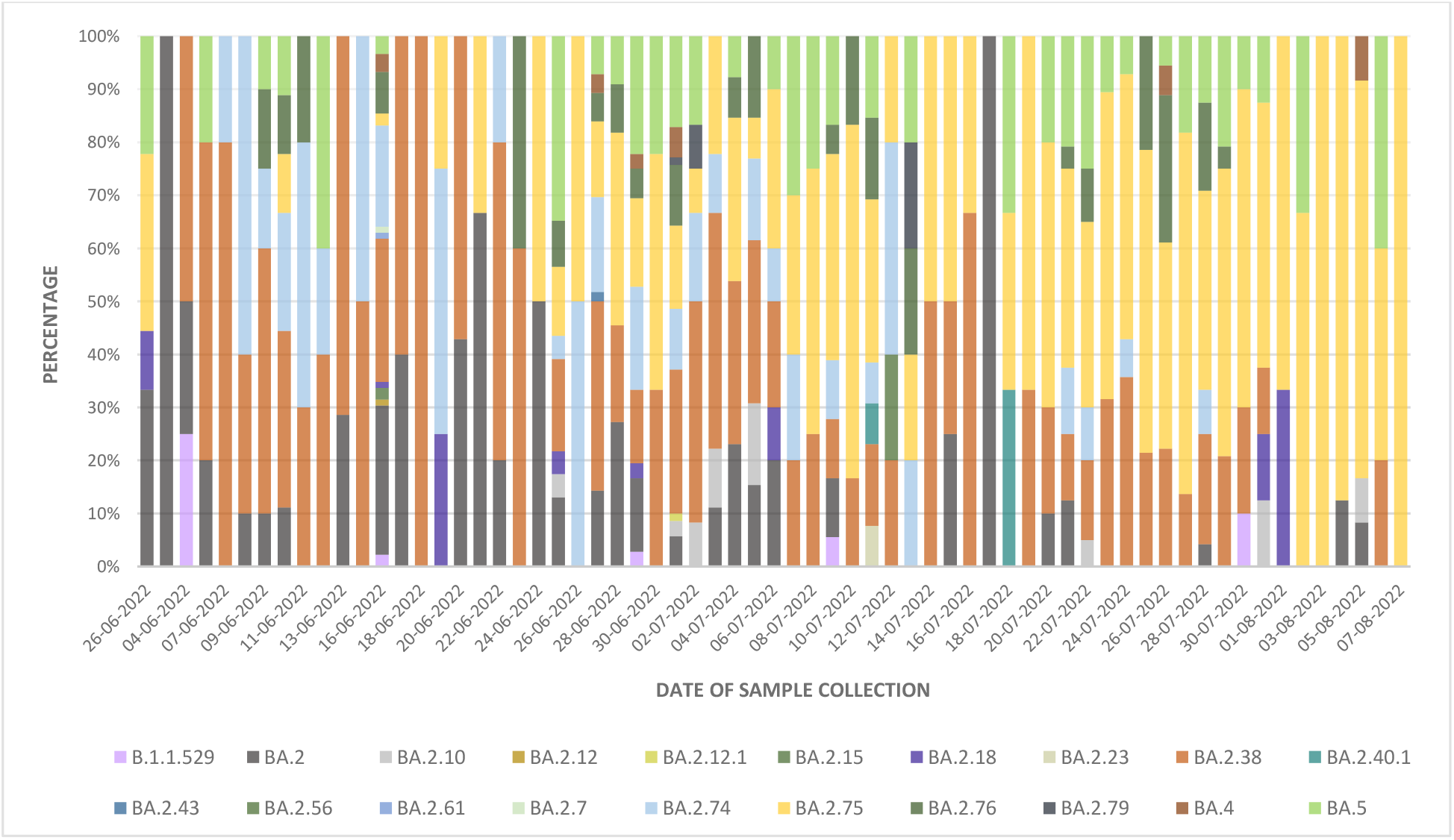
Distribution of variants to date of sample collection (From 06^th^ June 2022 to 7^th^ August 2022)

### 3.1 Demographic characteristics of the BA.2 sublineage confirmed cases (BA.2.74, BA.2.75, and BA.2.76)

The demographic and epidemiological characteristics of 371 BA.2 sub-lineage (BA.2.74, BA.2.75 and BA.2.76) confirmed patients are summarised in **Table 3**. Of these 371 cases, 212 (57.14%) were male, and 159 (42.86%) were female. The median age of the cases is 36 years, and the age group 21-40 years were predominantly affected.

**Table 3:** Demographic characteristics of the emerging Omicron variants

| Characteristics | Number of cases (%) |  |  |  |
| --- | --- | --- | --- | --- |
|  | BA.2.74 | BA.2.75 | BA.2.76 | Total |
| **1. Age Group (in years)** |  |  |  |  |
| 0-10 | 03<br>(3.41%) | 04<br>(1.75%) | 04<br>(7.27%) | 11<br>(2.96%) |
| 11-20 | 02<br>(2.27%) | 15<br>(6.58%) | 03<br>(5.45%) | 20<br>(5.39%) |
| 21-30 | 20<br>(22.73%) | 65<br>(28.51%) | 08<br>(14.54%) | 93<br>(25.07%) |
| 31-40 | 21<br>(23.86%) | 51<br>(22.37%) | 14<br>(25.46%) | 86<br>(23.18%) |
| 41-50 | 15<br>(17.05%) | 32<br>(14.04%) | 14<br>(25.46%) | 61<br>(16.44%) |
| 51-60 | 11<br>(12.50%) | 24<br>(10.53%) | 02<br>(3.64%) | 37<br>(9.97%) |
| 61-70 | 11<br>(12.50%) | 20<br>(8.77%) | 04<br>(7.27%) | 35<br>(9.43%) |
| 71-80 | 04<br>(4.55%) | 11<br>(4.82%) | 06<br>(10.91%) | 21<br>(5.66%) |
| >80 | 01<br>(1.14%) | 06<br>(2.63%) | 00<br>(00%) | 07<br>(1.89%) |
| **2. Gender** |  |  |  |  |
| Male | 50<br>(56.82%) | 131<br>(57.46%) | 31<br>(56.36%) | 212<br>(57.14%) |
| Female | 38<br>(43.18%) | 97<br>(42.54%) | 24<br>(43.64%) | 159<br>(42.86%) |
| **3. Area of Residence** |  |  |  |  |
| Pune | 69<br>(78.40%) | 216<br>(94.74%) | 48<br>(87.27%) | 333<br>(89.76%) |
| Mumbai | 17<br>(19.32%) | 02<br>(0.88%) | 07<br>(12.73%) | 26<br>(7.01%) |
| Solapur | 0<br>(00%) | 10<br>(4.38%) | 0<br>(00%) | 10<br>(2.69%) |
| Latur | 01<br>(1.14%) | 0<br>(00%) | 0<br>(00%) | 01<br>(0.27%) |
| Sindhudurg | 01<br>(1.14%) | 0<br>(00%) | 0<br>(00%) | 01<br>(0.27%) |
| **4. Calendar week of sample collection** |  |  |  |  |
| 22<sup>nd</sup> week of the year 2022<br>(30<sup>th</sup> May to 5<sup>th</sup> June 2022) | 00<br>(00%) | 00<br>(00%) | 00<br>(00%) | 00<br>(00%) |
| 23<sup>rd</sup> week of the year 2022<br>(6<sup>th</sup> to 12<sup>th</sup> June 2022) | 18<br>(20.45%) | 01<br>(0.44%) | 06<br>(10.91%) | 25<br>(6.74%) |
| 24<sup>th</sup> week of the year 2022<br>(13<sup>th</sup> to 19<sup>th</sup> June 2022) | 20<br>(22.73%) | 03<br>(1.32%) | 07<br>(12.73%) | 30<br>(8.09%) |
| 25<sup>th</sup> week of the year 2022<br>(20<sup>th</sup> to 26<sup>th</sup> June 2022) | 03<br>(3.41%) | 09<br>(3.95%) | 04<br>(7.28%) | 16<br>(4.31%) |
| 26<sup>th</sup> week of the year 2022<br>(27<sup>th</sup> June to 3<sup>rd</sup> July 2022) | 28<br>(31.82%) | 36<br>(15.79%) | 14<br>(25.45%) | 78<br>(21.02%) |
| 27<sup>th</sup> week of the year 2022<br>(4<sup>th</sup> to 10<sup>th</sup> July 2022) | 07<br>(7.95%) | 26<br>(11.40%) | 05<br>(9.09%) | 38<br>(10.24%) |
| 28<sup>th</sup> week of the year 2022<br>(11<sup>th</sup> to 17<sup>th</sup> July 2022) | 04<br>(4.55%) | 12<br>(5.26%) | 03<br>(5.45%) | 19<br>(5.12%) |
| 29<sup>th</sup> week of the year 2022<br>(18<sup>th</sup> to 24<sup>th</sup> July 2022) | 06<br>(6.82%) | 42<br>(18.42%) | 03<br>(5.45%) | 51<br>(13.75%) |
| 30<sup>th</sup> week of the year 2022<br>(25<sup>th</sup> to 31<sup>st</sup> July 2022) | 02<br>(2.27%) | 62<br>(27.19%) | 13<br>(23.64%) | 77<br>(20.76%) |
| 31<sup>st</sup> week of the year 2022<br>(1<sup>st</sup> August to 7<sup>th</sup> August 2022) | 00<br>(0%) | 37<br>(16.23%) | 00<br>(0%) | 37<br>(9.97%) |

### 3.2 Clinical characteristics of the BA.2 sublineage cases (BA.2.74, BA.2.75 and BA.2.76)

Of the 371 cases, 228 (61.46%) could be contacted to obtain information regarding symptoms, hospitalization status, treatment and vaccination status. **Table 4** describes the clinical characteristics and vaccination status of the confirmed cases of BA.2.74, BA.2.75 and BA.2.76.

**Table 4:**
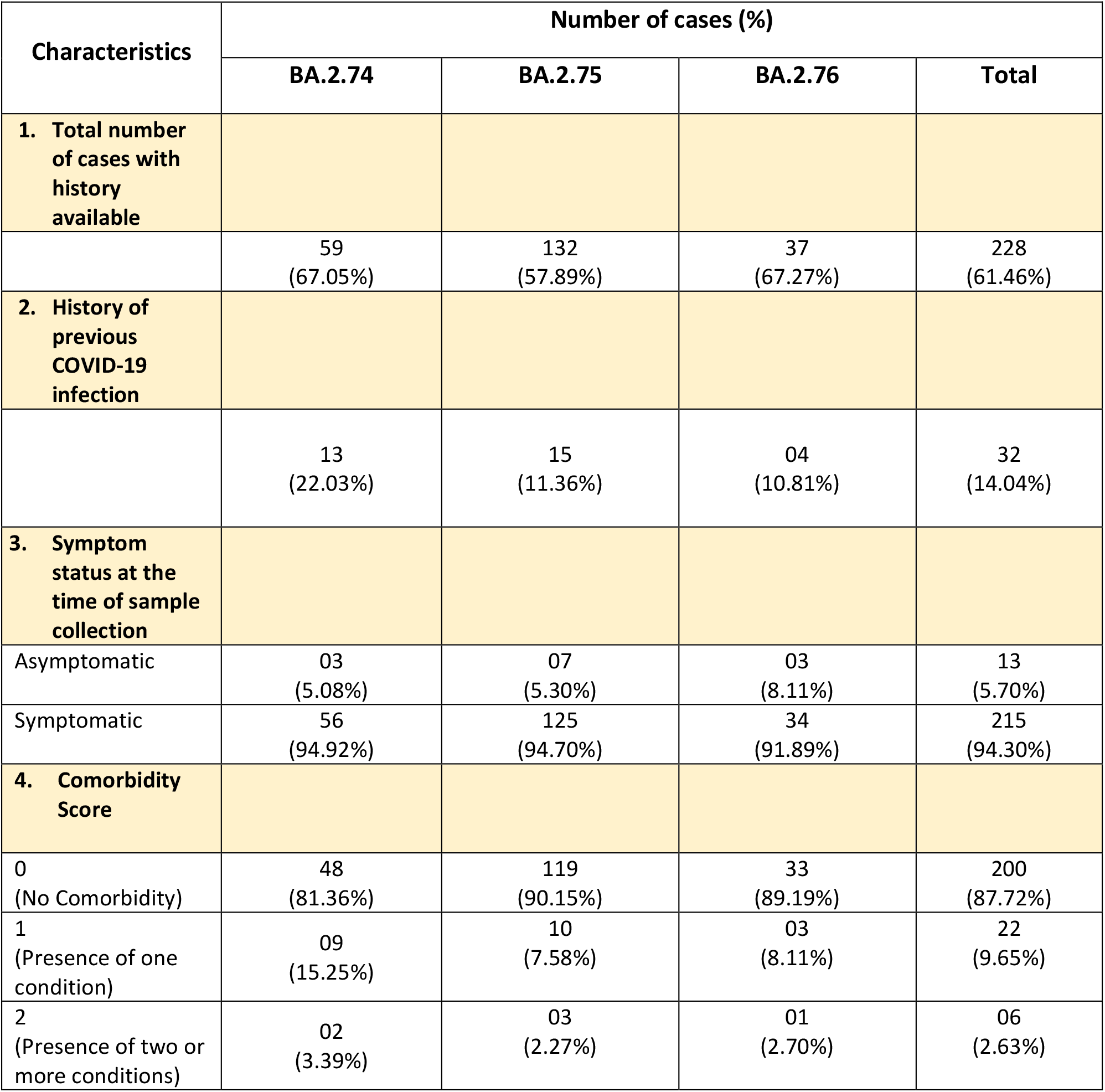
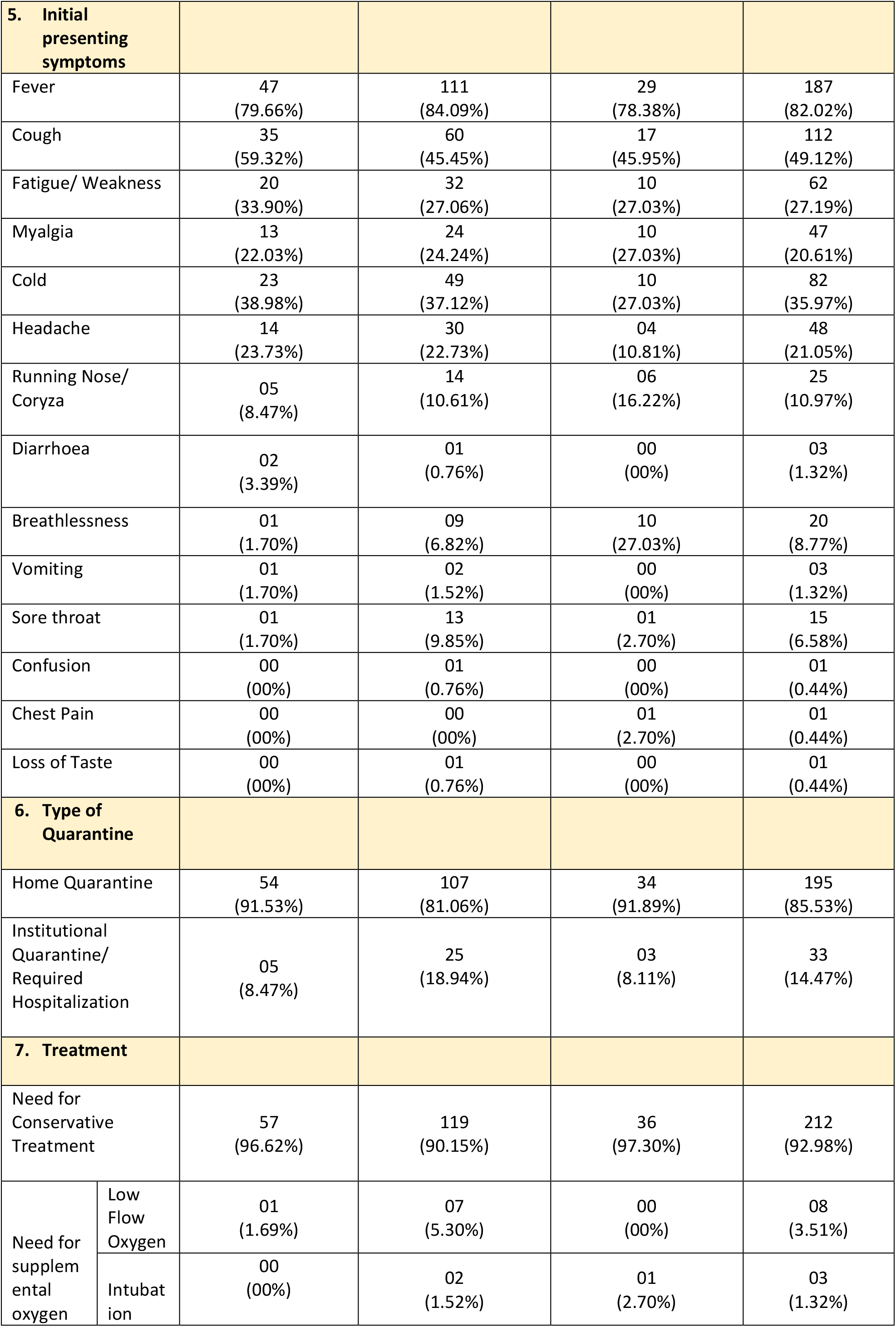
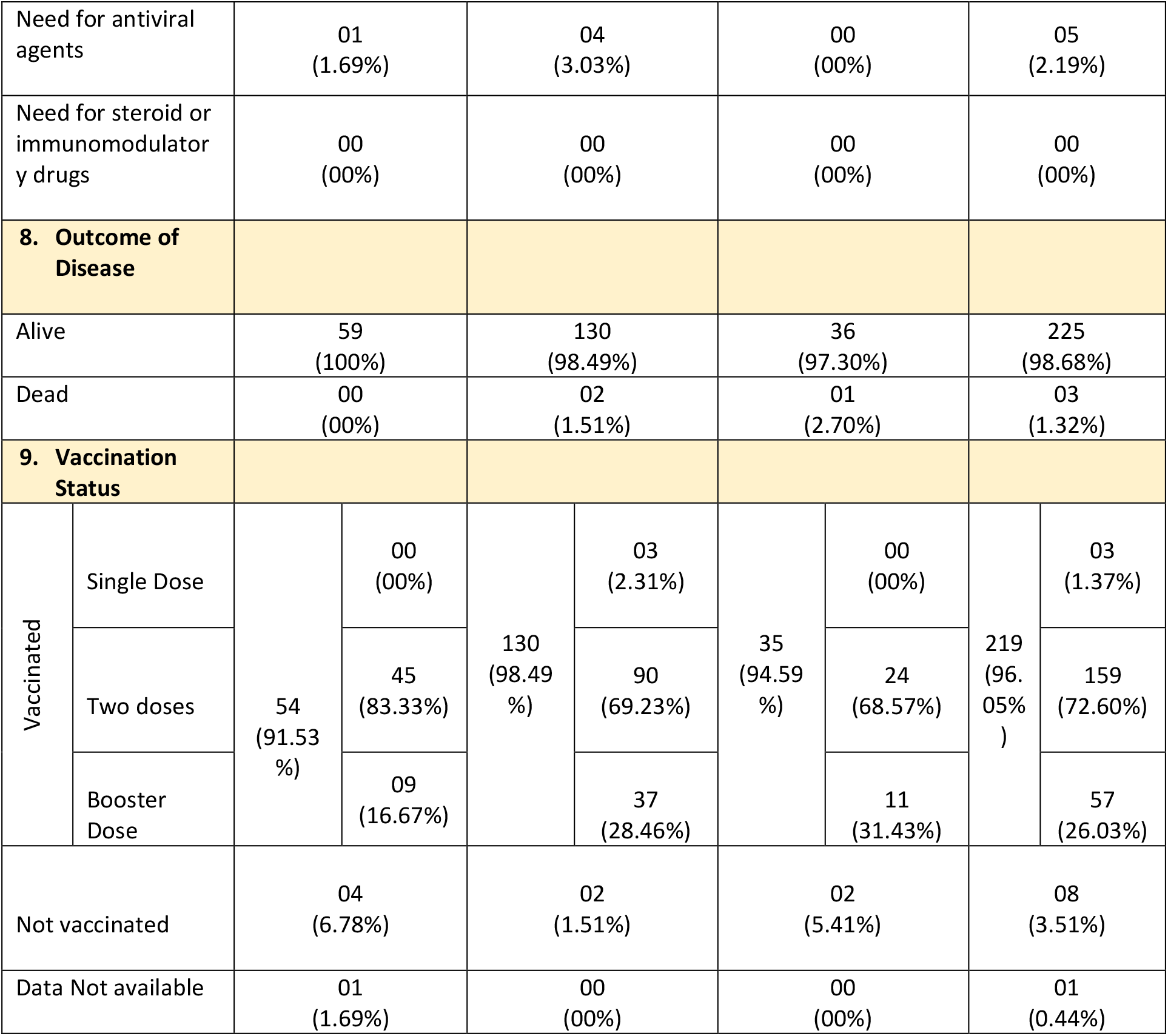
Clinical Characteristics and outcome of patients infected with emerging Omicron BA.2 sub-lineages

Among the 228 cases, 219 (96.05%) were vaccinated with at least a single dose of the COVID vaccine, and the remaining 08 (3.51%) were unvaccinated. Of the 219 vaccinated cases, 159 (72.60%) have received two doses of vaccine and 57 (26.03%) have received the precautionary dose. Three (1.37%) cases were vaccinated with a single dose only. All the unvaccinated cases were children less than 18 years.

Out of 228 cases, 215 (94.30%) developed mild symptoms, of which fever (82.02%) was the most common symptom, followed by cough (49.12%), cold (35.97%), fatigue (27.19%), headache (21.05%) and myalgia (20.61%). Rest 13 (5.70%) had an asymptomatic infection. There were 195 (85.53%) cases who recovered at home, and 33 (14.47%) required institutional quarantine. Of the 228 cases, 212 (92.98%) cases recovered with supportive treatment, 11 (4.83%) required supplemental oxygen therapy and 05 (2.19%) were given antiviral treatment. None were administered steroids, immunomodulatory drugs or monoclonal antibodies. There were 03 (1.32%) cases that progressed to severe disease that resulted in death, and the remaining 225 (98.68%) cases had good clinical outcomes.

## 4. DISCUSSION

One of the most characteristic features of the SARS-CoV-2 virus has been its rapid evolution during the pandemic. New variants have emerged from time to time with selective advantages, replacing the previously circulating variants and achieving global dominance. Barely weeks after the BA.2 lineage-driven surges globally, towards the third week of May 2022, India saw a surge in cases of COVID-19. (4) In the last 90 days, 219,503 whole genome sequences of SARS-CoV-2 have been deposited on GISAID from India. Of these, BA.2.38 (15%) is the predominant lineage, followed by BA.2.76 (14%), BA.2.75 (14%), BA.2 (9%), BA.5.2 (6%), and BA.2.74 (4%) **(Figure 2)**. (13) To date, 4,389 sequences of BA.2.76 (14), 3,321 of BA.2.75 (15), and 1,250 sequences of BA.2.74 (16) lineages have been deposited on GISAID. The apparent cumulative prevalence is less than 0.5% worldwide for all three lineages. In India, the prevalence of BA.2.76, BA.2.75 and BA.2.74 is 6%, 3% and 1%, respectively. Apart from India, BA.2.76 has been detected in 48 countries and 39 US states, BA.2.75 in 37 countries and 30 US states, and BA.2.74 in 32 countries and 23 US states. (14) (15) (16) When the world, particularly countries like South Africa, the United Kingdom, the United States, Germany, Portugal and Denmark, were experiencing the latest global outbreak driven by the BA.4 and BA.5 Omicron lineages (5), India, on the other hand, did not see an exponential increase in cases due to these two lineages. Based on the sequences deposited on GISAID, the BA.5 and BA.4 Omicron lineages continue to dominate globally, with a weekly prevalence of 69.6% and 11.8%, respectively. (17) However, it is interesting to note that, in India, the prevalence of BA.5 and BA.4 lineages is 9% and less than 0.5% among the sequences deposited on GISAID, respectively. (13) This probably could be as both Delta and BA.4/ BA.5 share the L452R mutation on the receptor-binding domain (RBD) of spike protein, the convalescent sera from Delta infection may contain L452R-specific neutralizing antibodies, which could have impaired the BA.4/ BA.5 transmission in India. (18) Also, BA.2.75 has shown 57-fold higher binding affinity to ACE2 receptors when compared with BA.5, accounting for its higher transmissibility. (19)

**Figure 2:**
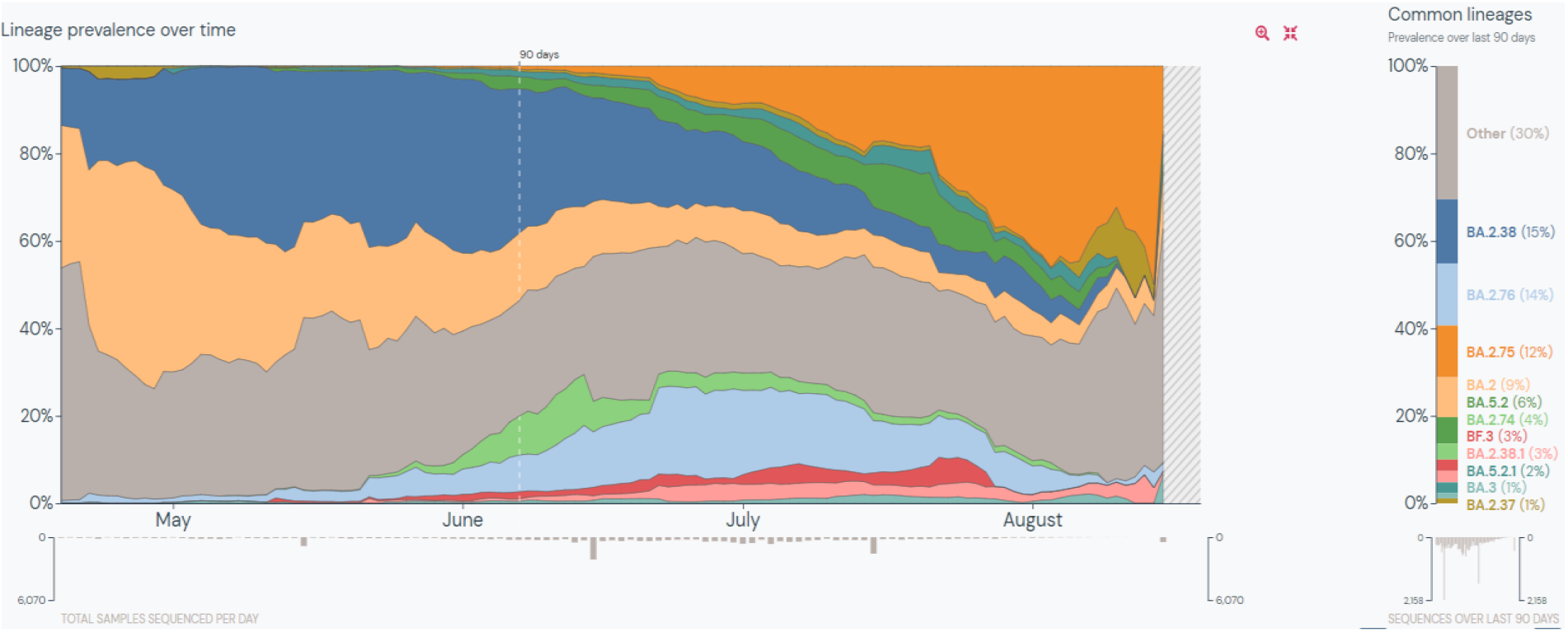
Lineage prevalence in India over the last 90 days (13)

The SARS-CoV-2 virus has evolved rapidly, adapting to its human hosts by developing mutations over time and resulting in the emergence of new variants. The Omicron subvariant BA.2.74 contains BA.2 mutations along with characteristic R346T and L452M mutations in its spike protein. (8) Similarly, the subvariant BA.2.76 contains BA.2 mutations with Y248N and R346T in the spike protein. (10) The subvariant BA.2.75, on the other hand, contains nine additional spike mutations compared to BA.2. The defining mutations include the BA.2 mutations with K147E, W152R, F157L, I210V, G257S in N-terminal domain and G339H, G446S, N460K, R493Q in the RBD region of spike protein **(Figure 3)**. (9) The effect of the mutations in the RBD region on the virus-host interactions is described in **Table 5**. (20) R493Q and N460K mutations increase the ACE-2 receptor affinity and surface RBD expression. (21) (22) On the other hand, mutations G446S and G339H decrease the ACE-2 affinity and RBD expression. Such adaptive mutations can alter the pathogenic potential of the virus.

**Table 5:** The defining spike mutations in the RBD region of three BA.2 sublineages and their effect on virus-host interaction (20)

| Variant | Mutations | Impact on |  |  |
| --- | --- | --- | --- | --- |
| | | ACE-2 affinity<br>$\Delta\log_{10} K_d$ | RBD expression<br>$\Delta\log$ (MFI) | Contact with ACE-2 receptor |
| BA.2.74 | R346T | 0.11 | 0.13 | No |
|  | L452M | 0.11 | 0.24 | No |
| BA.2.75 | G339H | -0.03 | -0.06 | No |
|  | G446S | -0.07 | -0.18 | Yes |
|  | N460K | 0.2 | 0.3 | No |
|  | R493Q | 1.07 | 0.04 | Yes |
| BA.2.76 | R346T | 0.11 | 0.13 | No |

**Figure 3:**
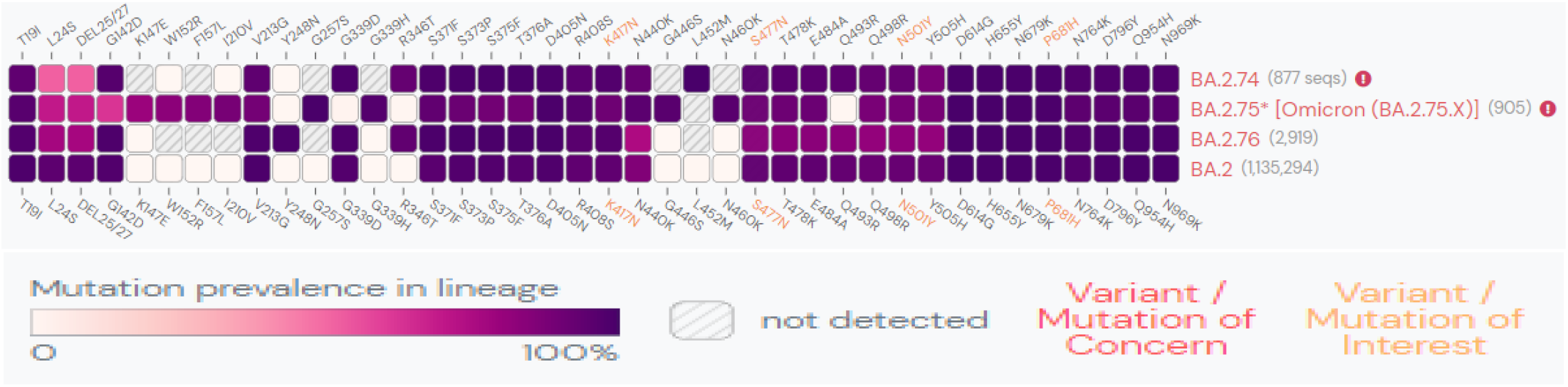
S-gene mutations in > 75% of global sequences for the three sublineages in the last 60 days (13)

**Table 6** compares the growth advantage of the common variants in India. These estimates reflect the advantage of the new variants compared to the cocirculating variants. The relative growth of a variant can be explained by three mechanisms: increased transmissibility, infectious duration, and immune evasion. The BA.2.75 sublineage has a relative growth advantage of 77% per week, with 0.08 as an assumed logistic growth rate per day. It has a 42% increase in transmissibility (23). The spike protein mutations D339H, G446S, N460K and R493Q allow BA.2.75 to escape neutralization by antibodies produced against different RBD epitopes in BA.2. (21) **Figure 4** shows the effect of antibodies elicited during pre-Omicron and Omicron BA.1 period on the BA.2.75 variant. The mutations in the RBD region confer an escape fraction of 0.45. (24) These features of BA.2.75 indicate that it might outcompete BA.4/ BA.5 and can become a potential risk to global health. Therefore, the spread and frequency of these sublineages in India and countries outside India require close monitoring through sustained genomic and clinical surveillance as they possess key mutations that are notable for their accelerated growth and extensive geographical distribution.

**Table 6:**
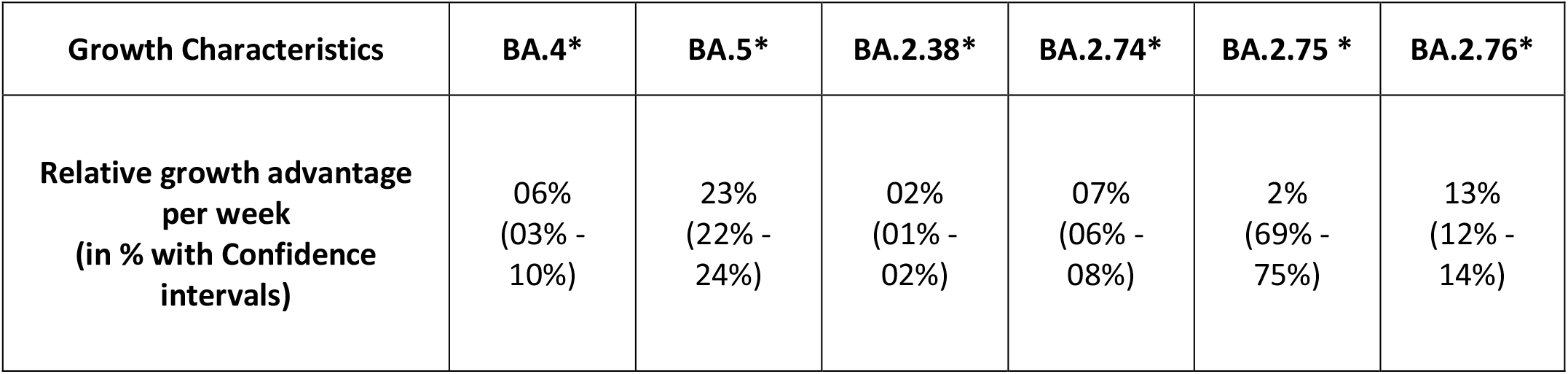
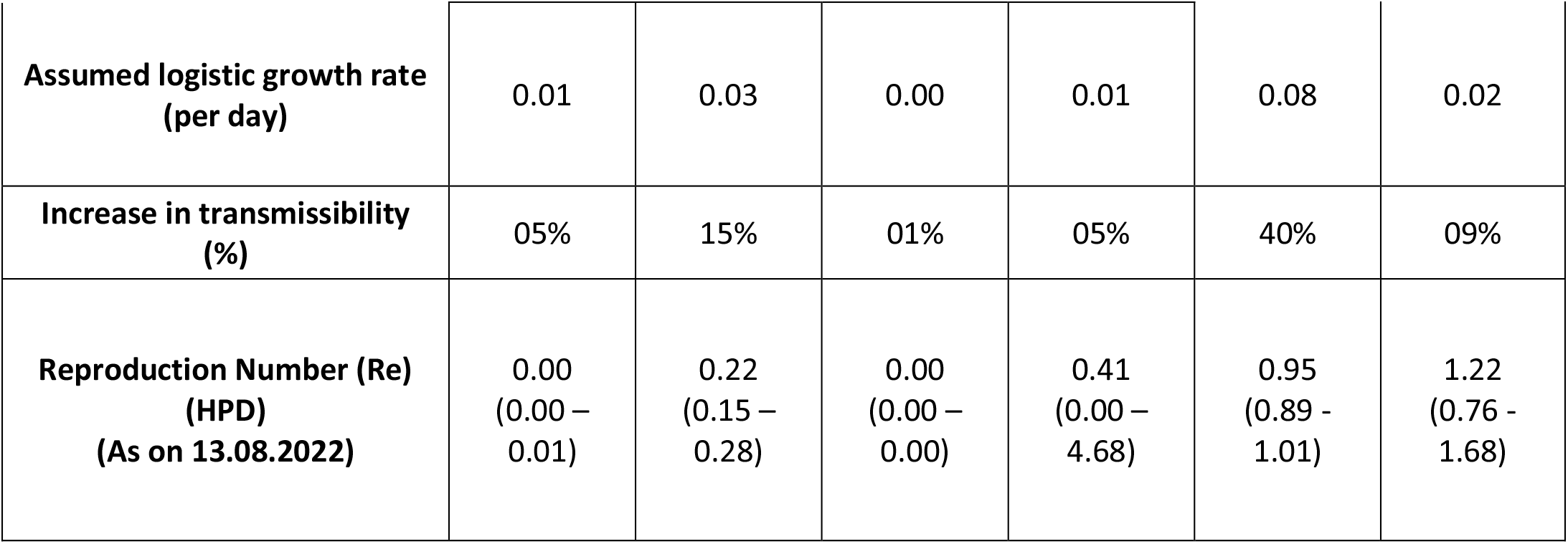
A comparative growth advantage of BA.4*, BA.5*, BA.2.38*, BA.2.74*, BA.2.75* and BA.2.76* in India (Data represented is as of 05-09-2022) (25)(26)(27)(22)(28)

| Growth Characteristics | BA.4* | BA.5* | BA.2.38* | BA.2.74* | BA.2.75 * | BA.2.76* |
| --- | --- | --- | --- | --- | --- | --- |
| Relative growth advantage per week (in % with Confidence intervals) | 06%<br>(03% - 10%) | 23%<br>(22% - 24%) | 02%<br>(01% - 02%) | 07%<br>(06% - 08%) | 2%<br>(69% - 75%) | 13%<br>(12% - 14%) |
| **Assumed logistic growth rate (per day)** | 0.01 | 0.03 | 0.00 | 0.01 | 0.08 | 0.02 |
| **Increase in transmissibility (%)** | 05% | 15% | 01% | 05% | 40% | 09% |
| **Reproduction Number (Re) (HPD) (As on 13.08.2022)** | 0.00<br>(0.00 – 0.01) | 0.22<br>(0.15 – 0.28) | 0.00<br>(0.00 – 0.00) | 0.41<br>(0.00 – 4.68) | 0.95<br>(0.89 - 1.01) | 1.22<br>(0.76 - 1.68) |

**Figure 4:**
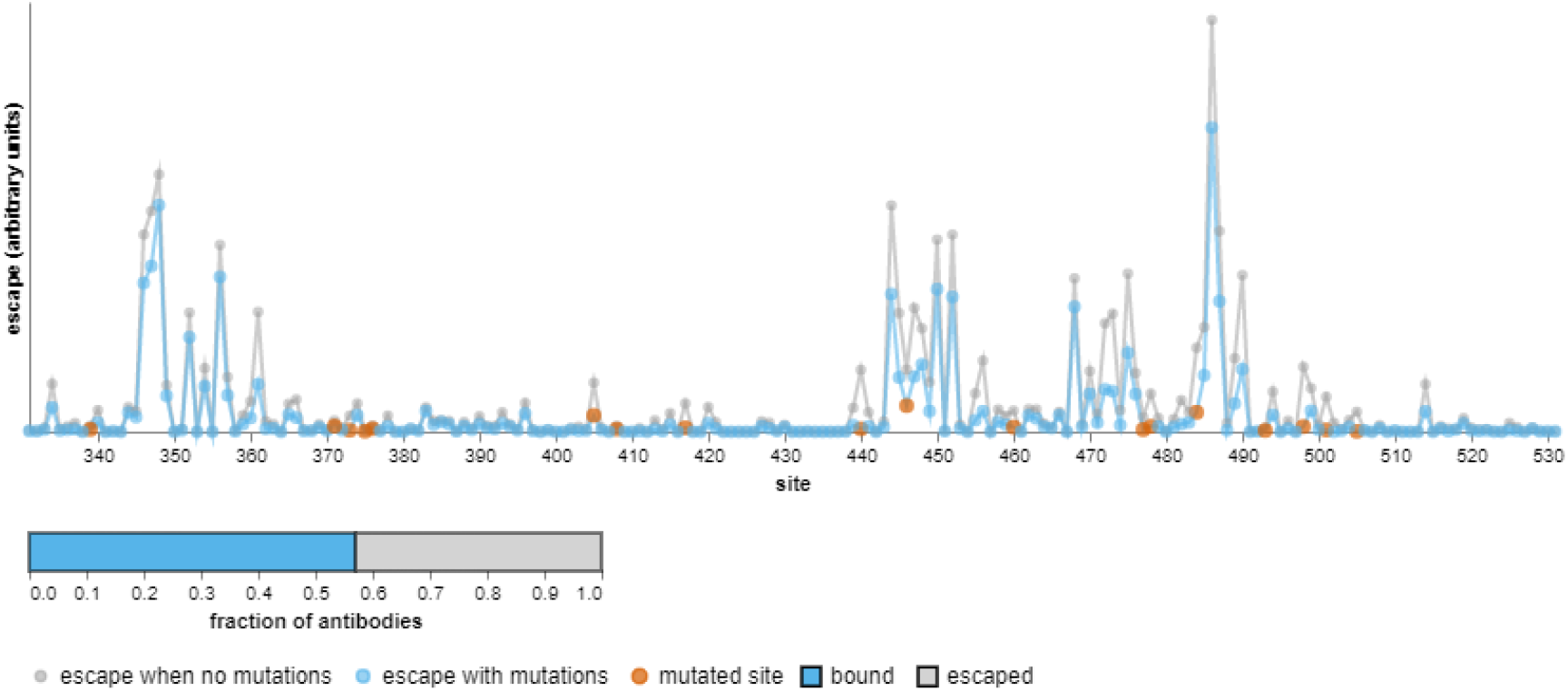
Effect of antibodies elicited during pre-Omicron SARS-CoV-2 and Omicron BA.1 period on neutralization of BA.2.75 variant* (24) *Escape fraction ranges from 0 to 1, where 0 means no escape and 1 means complete escape. Orange dots represent the site of mutation. Blue dots represent the escape in the presence of a mutation. Grey dots represent escape in the absence of a mutation.

The current study indicates that these BA.2 sublineages caused mild disease with reduced need for the hospital admission. In animal models, BA.2.75 replicated more efficiently in the lungs of hamsters than other Omicron variants causing focal pneumonia characterized by patchy inflammation in alveolar regions. These findings suggest that the Omicron subvariant BA.2.75 can cause severe respiratory disease and may affect the clinical outcome in infected humans. (29) However, it is still unclear to what extent the intrinsic virulence of the virus and the immunity due to vaccination or previous infections could have contributed to mild disease in India. Further, data from other clinical settings will be essential to assess the behaviour of these BA.2 sublineages in countries with different levels of previous infections and vaccination.

The fact that subvariant BA.2.75 contains mutations greater than BA.2 and BA.4/BA.5 raises concern regarding the possibility that it might have significantly reduced sensitivity to therapeutic monoclonal antibodies and antibodies developed by vaccination/natural infection. It is important to note that these subvariants have emerged when the world is about to achieve global immunity against the SARS-CoV-2 virus through various vaccines available for COVID-19. India has conducted 200 crore vaccinations and has vaccinated 73.02% of its population with at least one dose of vaccine and 66.85% of the population with two doses of vaccine. Around 7.67% of the total population has received the precautionary dose. (30) Various studies on the evasion of neutralizing antibodies have found that the Omicron subvariant BA.2.75, which has a local growth advantage in India, is 1.8 and 1.1 times more resistant to sera from vaccinated individuals than BA.2 and BA.2.12.1, respectively. However, it is 0.6-times more sensitive than BA.4/BA.5. It has increased resistance to class 1 and 3 monoclonal antibodies but is sensitive to Class 2 monoclonal antibodies. This increased resistance may be due to the spike protein’s G446S and R460K mutations. It is also 3.7 times more resistant to Bebtelovimab, the only monoclonal antibody potent against all Omicron subvariants. (21) Another study in a small sample of plasma from post-vaccination Delta infection shows that the BA.2.75 subvariant is more immune evasive than the BA.4 / BA.5 lineages in the Delta-stimulated immune background, which probably may explain why BA.2.75 has a growth advantage over BA.4/BA.5 in India. (18) Also, a study found that BA.2.75 has a higher resistance to BA.5 induced immunity. This property may make BA.2.75 variant spread efficiently in areas where BA.5 has been widely circulating. (31) Close monitoring and following these variants effectively and investigating their development as early as possible is crucial.

## 5. CONCLUSION

To conclude, this study provides essential and early evidence of the severity of the disease in patients infected with BA.2.74, BA.2.75 and BA.2.76 sublineages. Currently, there is no evidence of an increased risk of hospital admission or severe disease due to these sublineages in India. Despite these second-generation variants being detected only recently, they have the potential to be successfully transmitted across several countries due to the presence of critical mutations and significant growth advantages. The ability of the SARS-CoV-2 virus to evolve continuously and achieve increased transmission and immune evasion reinforces the importance of vaccination and sustained epidemiological surveillance to detect the emerging new variants.

## Data Availability

All data produced in the present work are contained in the manuscript

## ACKNOWLEDGEMENT

We acknowledge and thank Mrs Poonam Pacharne, Mr Vishal Rajput and Mrs Reena Katke for the technical help during the study.

